# COVID-19 prevention with alternate vaccine boosting frequencies for multiple-sclerosis patients undergoing therapy with beta-interferon, dimethyl fumarate, natalizumab, or teriflunomide

**DOI:** 10.1101/2024.09.19.24313891

**Authors:** Alex Dornburg, Hayley B. Hassler, Jeffrey P. Townsend

**Author notes:** Corresponding Author: Jeffrey P. Townsend PhD, 135 College St, Suite 200 #222, New Haven, CT 06510-2483.

## Abstract

Multiple-sclerosis patients undergoing treatment with disease-modifying therapies exhibit diverse immune responses to COVID-19 vaccinations. However, guidance on how specific treatments influence infection risks and optimal vaccination schedules remains limited. This study integrates data on vaccine-induced and infection-derived antibody responses to predict cumulative probabilities of breakthrough infections in untreated multiple-sclerosis patients and those treated with interferon, dimethyl fumarate, natalizumab, or teriflunomide. Using antibody dynamics and augmented logistic regression models, we evaluated the effectiveness of different Pfizer-BioNTech BNT162b2 booster schedules.

Our findings reveal that annual boosters effectively reduce infection risks for untreated multiple-sclerosis patients, lowering their cumulative risk by more than half over two years. Among treated patients, booster vaccinations generally provide protection comparable to that of untreated patients, although treatment-specific variations in immunity are evident. For patients on interferon, annual boosters yield an even greater reduction in risk. Patients treated with dimethyl fumarate or natalizumab benefit significantly from boosters, though they experience moderately higher risks compared to untreated patients.

This study underscores the importance of tailored booster schedules for MS patients, taking into account disease-modifying-therapy-specific effects on immunity. Our analysis provides actionable insights for mitigating SARS-CoV-2 risks in this vulnerable population until broader long-term infection data are available. These findings aim to guide clinicians in optimizing care for multiple-sclerosis patients in the context of ongoing COVID-19 vaccination strategies.

**Practice Points:**

1. **Tailor Booster Schedules:** Annual COVID-19 boosters are recommended for untreated MS patients and those on interferon, dimethyl fumarate, natalizumab, or teriflunomide; Bi-annual boosters would further reduce infection risk.
2. **Address Risks for Immunosuppressed Patients:** For patients on highly immunosuppressive treatments (e.g., fingolimod, ocrelizumab, rituximab), recognize diminished vaccine efficacy and consider supplemental measures for risk mitigation.
3. **Promote Booster Adherence and Education:** Encourage timely booster adherence and educate patients on the benefits of tailored vaccination schedules, while keeping in mind the potential antiviral properties of specific therapies.

## Introduction

COVID-19 vaccination strategies have primarily focused on identifying optimal booster frequencies that sustain immunity within the general population ^1,2^. For patients with multiple sclerosis (MS) undergoing disease-modifying therapies (DMTs), booster vaccination may be an essential tool for safe care. However, the impact of DMTs on the immune system varies significantly; some therapies appear to permit a relatively normal vaccine response, while others can severely impair the ability to respond to vaccinations ^3^. Data on antibody levels following COVID-19 vaccination justify a concern that atypical responses to DMTs might entail distinct booster schedules to achieve sufficient protection ^4,5^. Antibody responses are likely to vary depending on the specific DMT regimen ^6,7^, underscoring the need for specific guidance regarding infection risks associated with alternate frequencies of boosting ^8^.

## Methods

Anti-S antibody levels provide an accurate correlate of protection against infection by the SARS-CoV-2 coronavirus ^9^, therefore we quantified risks of infection on the basis of antibody levels. We conducted a literature search for anti-S antibody levels in individuals with MS vaccinated with Pfizer-BioNTech BNT162b2. We obtained data from MS patients undergoing treatment with interferon (*n* = 135), dimethyl fumarate (*n* = 161), natalizumab (*n* = 74), or teriflunomide (*n* = 56), from a group with untreated MS (*n* = 205), and from a control group without MS (*n* = 63) ^10^. Over the time points used in our analysis (4 weeks post original vaccination to 4 weeks post booster vaccination), the sampled population was 73–74% female, 48–49 years of age on average, and maintained a consistent Expanded Disability Status Scale of 2. To place antibody data into a comparative framework for analyses, we scaled antibody levels (Idda et al. 2022, Table 4) ^11^ by imputing the control group peak relative to MS patients and normalizing these values relative to the peak antibody level expected following BNT162b2 booster vaccination ^12^. There is currently no long-term antibody waning data on DMTs following vaccination. However, short-term data on the above therapies suggest no difference in waning between DMT patients and a control group or non-MS patients ^13^. To predict the trajectory of antibody decline in these cohorts over longer terms, we incorporated longitudinal waning data for anti-N and anti-S IgG antibodies so as to obtain adequate phylogenetic representation of waning of antibodies responsive to six coronaviruses—HCoV-OC43, HCoV-NL63, HCoV-229E, SARS-CoV-1, SARS-CoV-2, and MERS. As in Townsend et al. ^14^, we utilized a linear model to relate anti-N and anti-S antibody levels—an appropriate model to apply because their decline is strongly correlated ^15^. We subjected the antibody waning data to an ancestral and descendant states analysis ^14,16^ to infer a baseline antibody level and an augmented exponential waning parameter for SARS-CoV-2. Ancestral and descendant states analysis is a well-established technique commonly employed in evolutionary biological research that analyzes the phylogenetic relationships and divergence times between lineages with a model of trait evolution to estimate unknown character states for past or current lineages ^17–19^. Extensive analyses have demonstrated that imputed antibody levels supplied by ancestral and descendent states analyses provide robust estimates of antibody waning profiles ^14,20^ following vaccination against SARS-CoV-2 that have been empirically validated ^21,22^. Antibody waning profiles for each cohort were generated based on cohort-specific peak antibody responses. To relate these antibody waning profiles to daily endemic infection probabilities, we fit a logistic infection function as in Townsend et al ^14^. Ancestral and descendent state analysis provided the slope and intercept parameters of the logistic function relating daily antibody level to daily infection probability. These infection probabilities in turn enabled calculation of the cumulative risks of breakthrough infection over time. We then compared these risks across booster schedules ^12^ with each booster conferring antibody responses as reported in Pitzalis et al. ^10^.

## Results

Annual booster vaccinations over a two-year period provided equivalent protection for individuals without MS and untreated patients with MS, reducing infection risk by more than half (28–29% risk without boosting versus 11% with annual boosting; **Figure 1**). Notably, MS patients receiving interferon therapy experienced an even greater benefit from annual boosters, with their risk of infection dropping to just 6%—a nearly four-fold reduction of the 23% risk without boosting. Patients on dimethyl fumarate, natalizumab, or teriflunomide faced a range of moderately elevated infection risks relative to untreated patients; without boosters, nearly one-third of patients undergoing dimethyl fumarate or natalizumab treatment could become infected over two years. Annual boosting reduced this risk by half, resulting in a 15% risk over the same period. Patients treated with teriflunomide would be more vulnerable to infection relative to untreated MS patients (22% versus 11%) based on their antibody response. However, this prediction regarding teriflunomide is complicated by its unmodeled antiviral properties ^23^.

**Figure 1.**
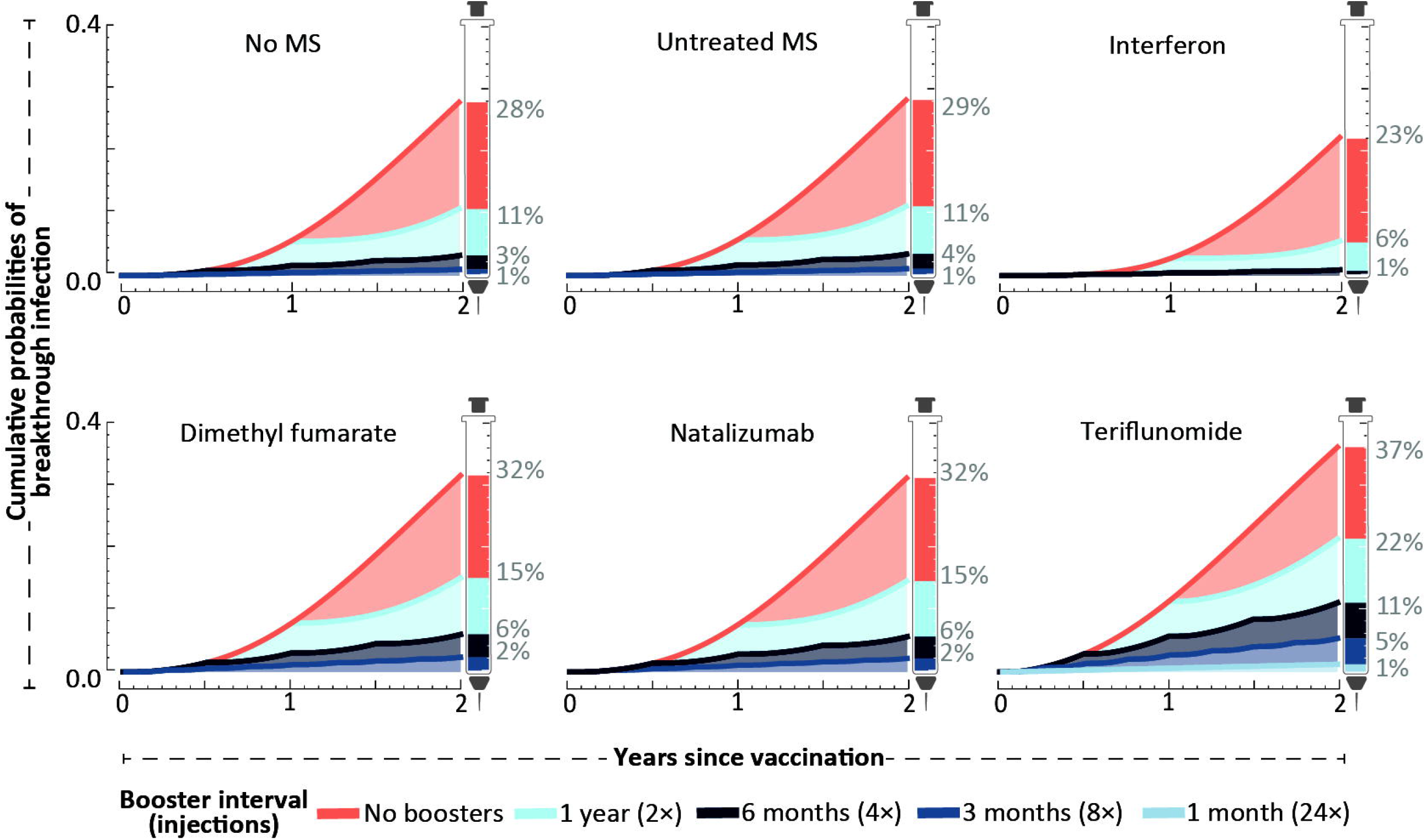
Cumulative probabilities of breakthrough infection over a two-year time span for updated BNT162b2 booster doses after primary vaccination, over intervals of one, three, six, or twelve months (colored bands). The analysis compares the general population, untreated MS patients (*n* = 205), and MS patients receiving one of four disease-modifying therapies: interferon (*n* = 135), dimethyl fumarate (*n* = 161), natalizumab (*n* = 74), and teriflunomide (*n* = 56) ^10^.

## Discussion

Our analysis indicates that patients undergoing several MS treatments exhibit risks of infection that are fairly similar to risks for the general population. We showed that frequent boosting in these populations correlates with better protection. Our analysis of antibody levels and infection probabilities indicates that patients undergoing treatment with interferon, dimethyl fumarate, natalizumab, and teriflunomide would benefit substantially from booster vaccination. MS patients on dimethyl fumarate or natalizumab therapies appear to be at moderately higher infection risk and may warrant additional attentiveness to timely adherence as well as consideration for more intensive vaccination schedules. Teriflunomide-treated patients appeared to face the highest risk; a bi-annual booster schedule might be necessary for patients on teriflunomide to achieve comparable protection against infection.

Our analysis is based solely on the effects of these treatments on antibody levels, and the consequences of those antibody responses to vaccination on infection probability. Some DMTs increase or diminish the antibody response. For instance, interferon increases antibody response to booster vaccination and therefore in our analysis diminishes risk of infection for all booster schedules. On the other hand, some interferon therapies have been linked to increased risk of leukopenia ^24–26^, which in turn could increase risk of some infections. Nevertheless, studies of the effect of interferon treatment in MS have demonstrated—consistent with our result—that patients are not subject to a generalized increase in risk of viral infection ^25^.

Conversely, Teriflunomide has been shown to inhibit activated T and B cells ^25^, and decreases antibody response to booster vaccination ^10^. This lower initial antibody response increases risk of infection for all booster schedules. However, studies of teriflunomide have also demonstrated that this therapy has antiviral properties against SARS-CoV-2 ^23^. These unmodeled antiviral properties may lead to protection from infection, and therefore justify some concern that our results for teriflunomide may overstate the actual risk of infection at each booster vaccination frequency.

More potently immunosuppressive DMTs exist, including fingolimod, ocrelizumab, and rituximab. These DMTs can reduce the abundance of lymphocytes and alter various B-cell and T-cell traits, thereby impairing the ability to mount an effective immune response post-vaccination. Predicting the effects of these more immunosuppressive therapies poses even greater challenges. The limited data available for our analyses of these treatments (*n* < 10; ^10^) indicate that in recently vaccinated MS patients treated with these therapies, the levels of antibodies are typically lower than baseline unvaccinated antibody levels in non-MS populations. Our rates of antibody decline and probabilities of infection are based on those non-MS populations, in whom antibody levels do not decline below this baseline. Consequently, there is no empirical basis enabling our projection of antibody levels subsequent to booster vaccination, nor to associate these low levels of antibody with probabilities of infection. In this context, ascertaining the effect of boosting is highly speculative. However, with such low levels of antibodies, risk of infection is high. Patients on fingolimod and ocrelizumab have been shown to have a reduced response to vaccination against SARS-CoV-2 ^10,27,28^, and research on cancer patients treated with rituximab has indicated that even monthly boosters would fail to confer substantial protection ^29^.

Our study uses infection data from fully endemic coronaviruses for long-term predictions, which means it is based on responses to evolving viruses and therefore is appropriate to scenarios involving regularly updated vaccines that target predominant strains. For the same reason, our analysis accounts for waning vaccine efficacy due to antigenic evolution. The infection probabilities we calculated are not based on early trial data, which has limitations for long-term predictions because the immune systems of early trial participants were largely naive to the virus, requiring significant immunological adaptation to develop effective cellular immunity. Our study does not account for antigenic changes occurring between vaccine production and deployment, which could reduce booster efficacy. Regardless, our findings provide crucial guidance for mitigating SARS-CoV-2 infections in MS patients undergoing various DMTs until extensive longer-term infection data becomes available in this distinct patient population.

## Acknowledgements

We thank members of the Townsend and Dornburg lab groups for helpful comments and support.

## Data Availability Statement

All data, code, and a detailed readme have been archived on Zenodo and are currently available: https://zenodo.org/doi/10.5281/zenodo.13384814.

## Prior Presentation

This work has not been read or exhibited at a meeting.

